# Network meta-analysis combining survival and count outcome data: A simple frequentist approach

**DOI:** 10.1101/2025.01.23.25321051

**Authors:** Hisashi Noma, Kazushi Maruo

## Abstract

Network meta-analysis for survival outcome data often involves several studies only reported dichotomized outcomes (i.e., the numbers of events and sample sizes of individual arms). To avoid the reporting biases via eliminating these studies in the syntesis analyses, Woods et al. (2010; *BMC Med Res Methodol* 10:54) proposed a Bayesian approach to combine the survival and dichotomized outcome data using hierarchical models. However, the Bayesian methods require complicated computations involving the Markov Chain Monte Carlo and the technical aspects are generally difficult to handle by non-statisticians (e.g., the convergence diagnostics). Besides, frequentist approaches have been alternative standard methods for statistical analyses of network meta-analysis. The methodology has been well established, and several effective software packages have been developed (e.g., `netmeta` package in R). In this article, we propose a simple method to synthesis the survival and dichotomized outcome data within the frequentist framework using the contrast-based models. We also provide a R package `survNMA` (https://doi.org/10.32614/CRAN.package.survNMA) to calculate hazard ratio statistics from the dichotomized outcome data that can be directly used for implementing general analyses of network meta-analysis using `netmeta` package of R.

## 1. Introduction

Network meta-analysis for survival outcome data often involves several studies only reported dichotomized outcomes (i.e., the numbers of events and sample sizes of individual arms). To avoid the reporting biases via eliminating these studies in the synthesis analyses, Woods et al. ^1^ proposed a Bayesian approach to combine the survival and dichotomized outcome data using hierarchical models. However, the Bayesian methods require complicated computations involving the Markov Chain Monte Carlo ^2,3^ and the technical aspects are generally difficult to handle by non-statisticians (e.g., the convergence diagnostics). Besides, frequentist approaches have been alternative standard methods for statistical analyses of network meta-analysis ^4-6^. The methodology has been well established, and several effective software packages have been developed (e.g., `network` ^7^ in Stata, `netmeta` ^8^ and `NMA` ^9^ packages in R). In this article, we propose a simple method to synthesis the survival and dichotomized outcome data within the frequentist framework of network meta-analysis using the contrast-based models. We also provide a R package `survNMA` (https://doi.org/10.32614/CRAN.package.survNMA) to calculate hazard ratio statistics that can be directly used for implementing general analyses of network meta-analysis using `netmeta` package ^8^ of R.

## 2. Methods

In meta-analyses of survival outcome data, hazard ratio has been widely adopted as the effect measure ^10^, which can be estimated the commonly used Cox regression model ^11^. Woods et al. ^1^ proposed Bayesian hierarchical modelling methods to model the hazard ratio estimates and the dichotomous outcome data simultaneously. Besides, the contrast-based approach of frequentist framework for network meta-analysis requires summary statistics of contrast measures (e.g., hazard ratio, risk ratio) for the pooling analysis ^12,13^.

In this case, the log hazard ratio estimates and their standard error estimates are required for all eligible trials. If valid log hazard ratio estimates and their standard error estimates are calculated from the dichotomized outcome data, the pooling analyses can be simply conducted using existing standard methods. Salika et al. ^14^ investigated comparative effectiveness of several methods for computing the log hazard ratio estimates and their standard error estimates from dichotomized data through a large dataset by the Cochrane Database of Systematic Reviews, and concluded the estimating method using the complementary log-log link was favorable among the candidate methods.

Suppose *d*_1_, *d*_0_ represent the numbers of events, and *n*_1_, *n*_0_ represent the sample sizes of the corresponding two groups that the log hazard ratio should be estimated. Also, denote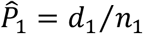 and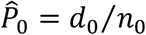 as the cumulative event proportions of the two groups. Then, the log hazard ratio estimator ^14^ is provided as

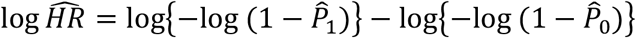

and the corresponding variance estimator ^14^ is

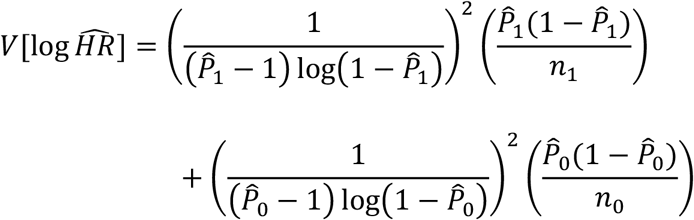

The log hazard ratio estimator is provided via the use of the complementary log-log link for a link function in the discrete-time hazards models ^15,16^, and the asymptotic normality is fulfilled for the summary statistic. Thus, the resultant summary statistics can simply be added to the remaining data that are obtained from the other studies that reported hazard ratio estimates. The contrast-based network meta-analysis methods ^4^ can be simply applied to the combined dataset, and then, valid pooling analyses can be conducted. The advantage of this approach is that all of the standard analysis tools for network meta-analysis can be directly applied to the combined dataset, e.g., pooling analyses, graphical tools, and heterogeneity and inconsistency assessments. As shown in the following section, all of the generic functions of `netmeta` package ^8^ in R can be applied to the combined dataset straightforwardly.

## 3. Implementation of the proposed method using “survNMA” package

### 3.1 Installation of “survNMA” package

The `survNMA` package can be installed from CRAN by the following command:

~~~
>install.packages(“survNMA”)
~~~

### 3.2 Loading the package

After the installation, users can load the package via the following command.

~~~
>library(“survNMA”)
~~~

### 3.3 Preparing a summary survival outcome data

As like ordinary meta-analysis of survival outcome data, the log hazard ratio estimates and their standard error estimates are needed in the analyses using contrast-based models. If these statistics can be directly extracted from literatures or back-calculations from related statistics (e.g., confidence intervals or P-values; see Parmar et al. ^17^ for the details), they can be straightforwardly used for the synthesis analyses. However, in multi-arm trials, these statistics are possibly not reported for several contrasts of comparative groups. In Table 1, we present summary hazard ratio statistics of two clinical trials in the network meta-analysis of chronic obstructive pulmonary disease (COPD) provided in Woods et al. ^1^. For the second study, the hazard ratio estimate and the standard error estimate were not reported for the comparison Salmeterol vs. Fluticasone. However, the other treatment pairs, these summary statistics are available. In these cases, Woods et al. ^1^ provided an approximation method for calculating these summary statistics from the available data.

**Table 1.** Example summary hazard ratio dataset for network meta-analysis of COPD (cited from Woods et al. ^1^) ^†^.

| Study | Treatment 1 | Treatment 2 | N1 | N2 | $\log \widehat{HR}$ | $\sqrt{V[\log \widehat{HR}]}$ |
| --- | --- | --- | --- | --- | --- | --- |
| Burge et al. (2000) | Fluticasone | Placebo | 376 | 375 | -0.276 | 0.203 |
| Calverly et al. (2007) | SFC | Placebo | 1533 | 1524 | -0.209 | 0.098 |
|  | Salmeterol | Placebo | 1521 | 1524 | -0.154 | 0.096 |
|  | Fluticasone | Placebo | 1534 | 1524 | 0.055 | 0.092 |
|  | SFC | Salmeterol | 1533 | 1521 | -0.056 | 0.100 |
|  | SFC | Fluticasone | 1533 | 1534 | -0.264 | 0.096 |

<sup>†</sup> The log hazard ratio estimates and their standard error estimates of Salmeterol vs. Fluticasone for Calverly et al. (2007) were not reported.

Without loss of generality, we suppose there are three groups A, B and C for comparison, and the summary hazard ratio statistics are available for the contrasts B vs. A and C vs. A; however, those for the contrast B vs. C are not available. In such cases, the log hazard ratio estimate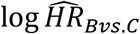 is calculable by

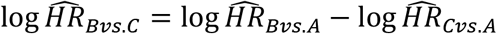

and the variance of 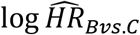is estimated by

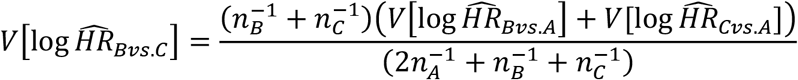

where *n*_*A*_, *n*_*B*_ and *n*_*C*_ are the sample sizes of the groups A, B and C. The standard error estimate can be easily calculated using the `calcse` function:

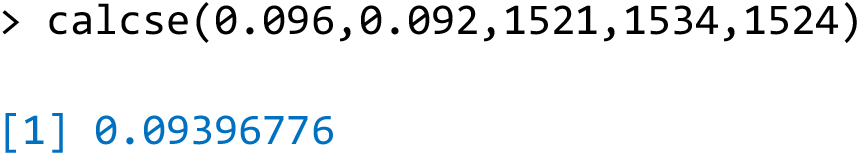

Using these outputs, users can make a summary dataset on R or other data editing software (e.g., Microsoft Excel); these datasets can be combined with the summary dataset created from dichotomized dataset as described in Section 3.5.

### 3.4 Calculation of the log hazard ratio estimates from dichotomized dataset

The log hazard ratio estimates and their standard error estimates can be calculated by the `pairwiseHR` function from dichotomized dataset. The `pairwiseHR` function simply requires the numbers of events `d` and the sample sizes `n` for all arms involved in the corresponding studies; the ID of studies `studlab` and treatment variable `treat` are also needed. We present an example dataset in Table 2; 3 clinical trials data that only reported dichotomized outcome data in the network meta-analysis of COPD ^1^ is presented.

**Table 2.** Example dichotomized dataset for network meta-analysis of COPD (cited from Woods et al. ^1^)^†^.

| Study | Treatment | d (death) | n | $\log \widehat{HR}$<br>(vs. Placebo) | $\sqrt{V[\log \widehat{HR}]}$ |
| --- | --- | --- | --- | --- | --- |
| Boyd et al. (1997) | Salmeterol | 1 | 229 | -0.009 | 1.414 |
|  | Placebo | 1 | 227 | — | — |
| TRISTAN (2003) | Fluticasone | 4 | 374 | -0.599 | 0.627 |
|  | Salmeterol | 3 | 372 | -0.883 | 0.690 |
|  | SFC | 2 | 358 | -1.251 | 0.802 |
|  | Placebo | 7 | 361 | — | — |
| Celli et al. (2003) | Salmeterol | 1 | 554 | -1.415 | 1.225 |
|  | Placebo | 2 | 270 | — | — |

<sup>†</sup> The log hazard ratio estimates and their standard error estimates were calculated using the `pairwiseHR` function.

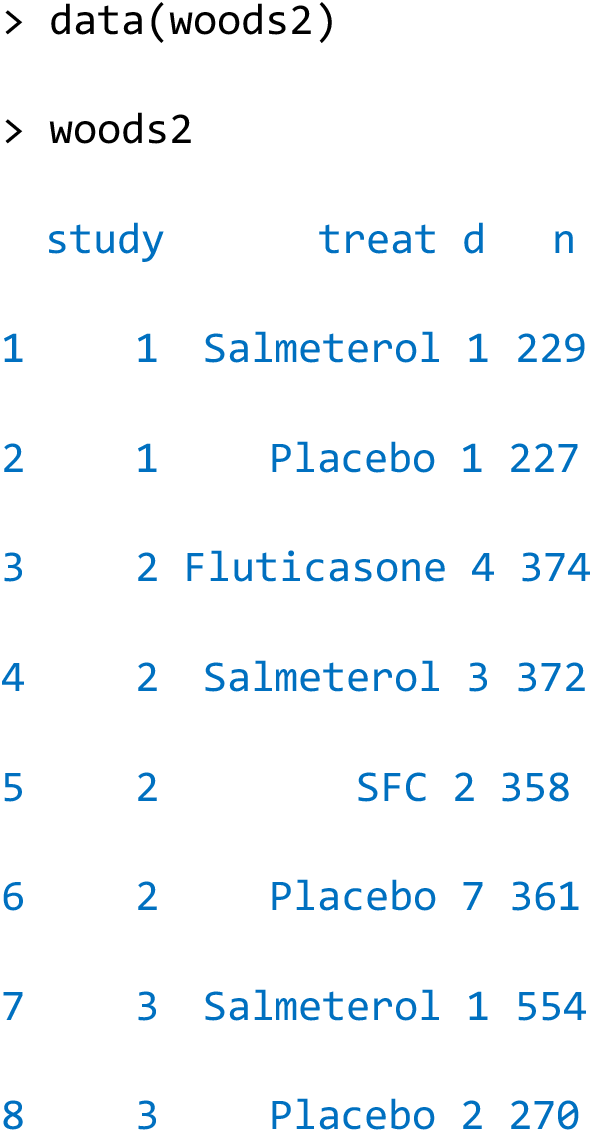

We can calculate the log hazard ratio estimates and their standard error estimates of these studies simultaneously by a simple command,

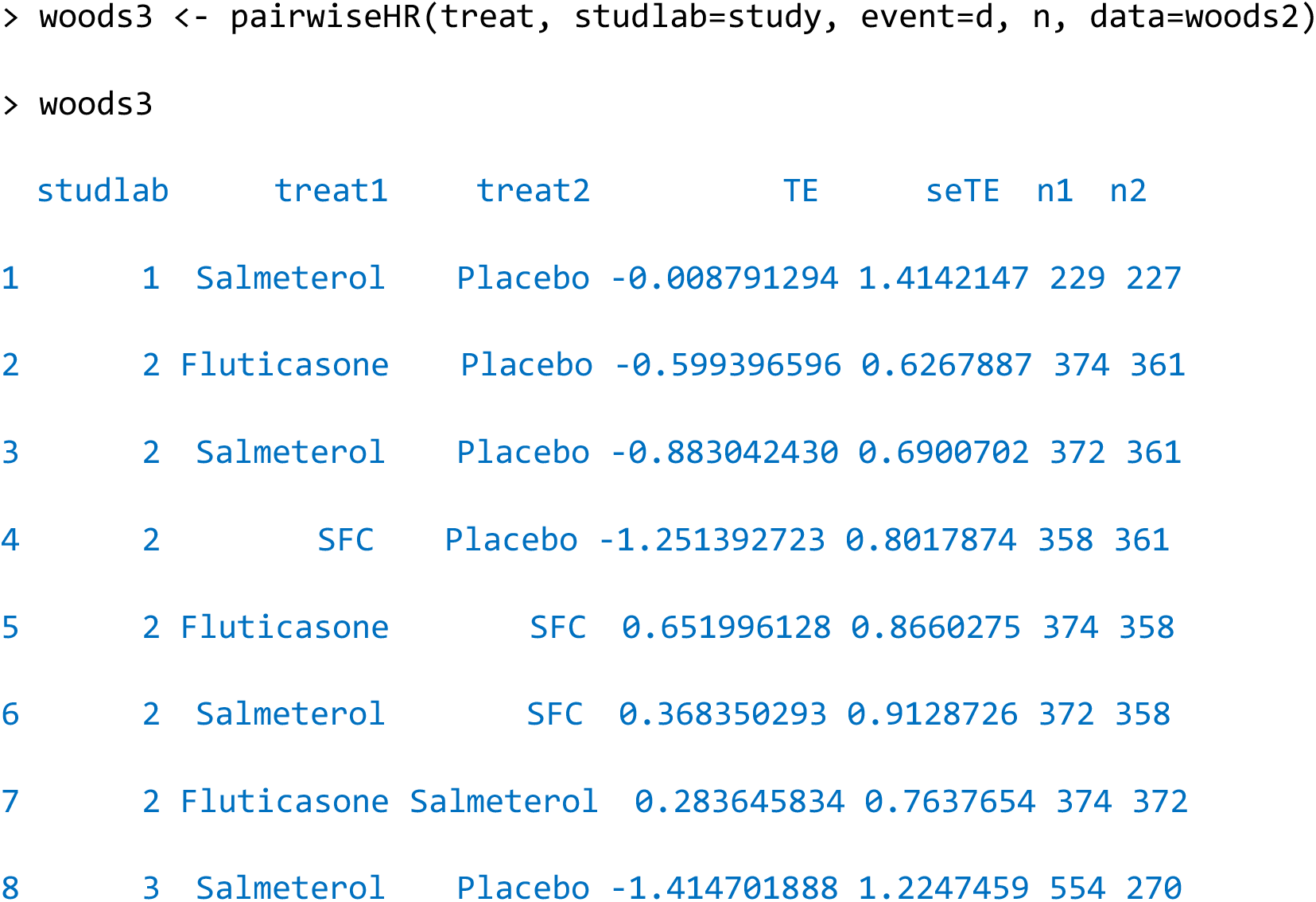

Using the output object, users can make a summary dataset that can be directly used for the network meta-analysis using the `netmeta` package.

### 3.5 Combining the dataset objects

The two dataset objects created by the survival and dichotomized outcome data (described in Sections 3.3 and 3.4) can be combined by `combine2` function.

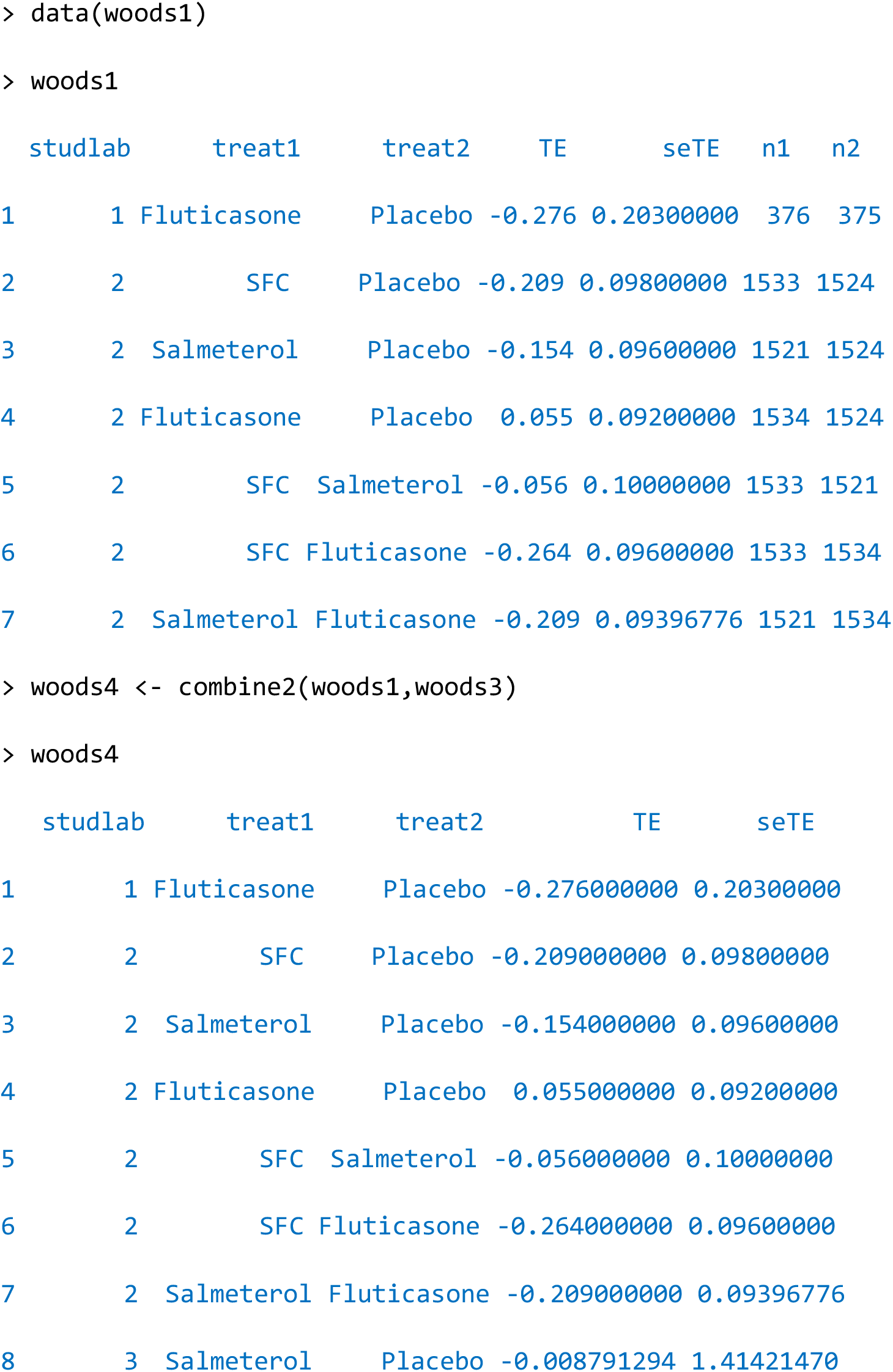
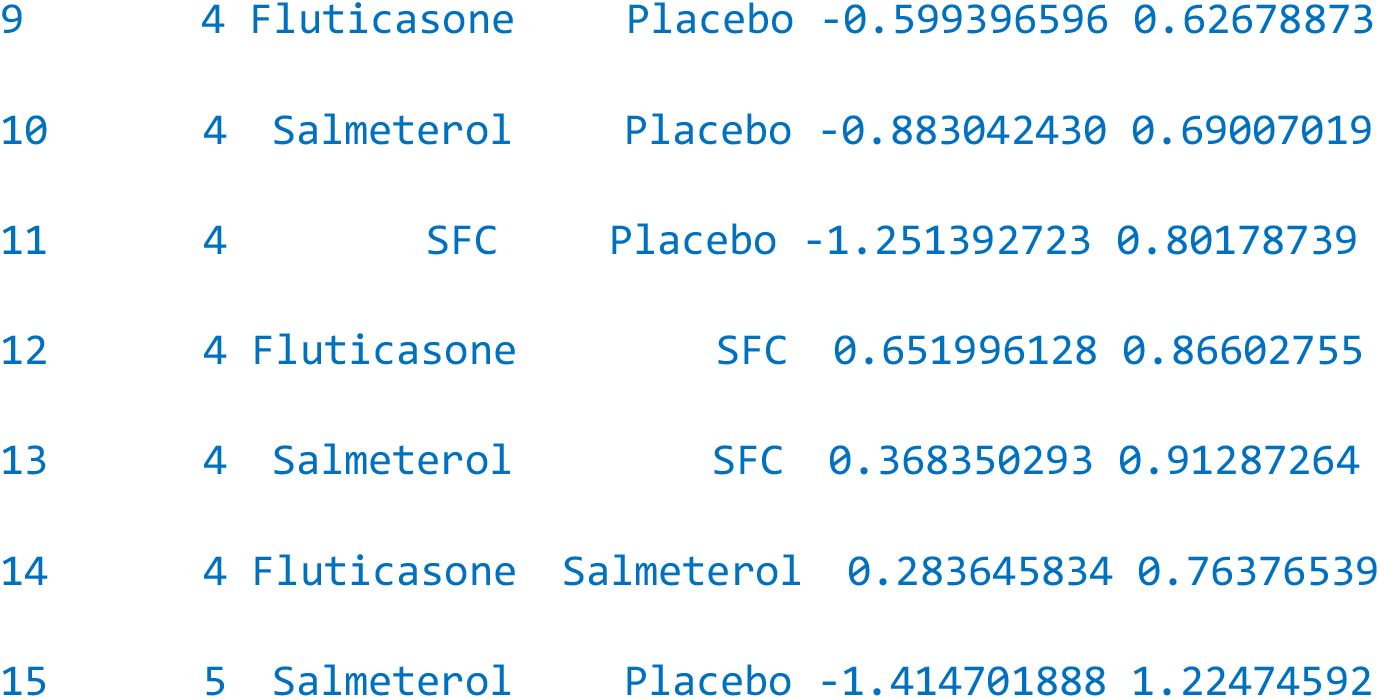

The output object can directly be used for general analyses of network meta-analysis using `netmeta` package.

### 3.6 Network meta-analysis for the combined dataset

The network meta-analysis can be conducted by simply applying existing packages for network meta-analysis to the combined dataset. For example, the pooling analysis can be simply performed by `netmeta` function in `netmeta` package ^8^,

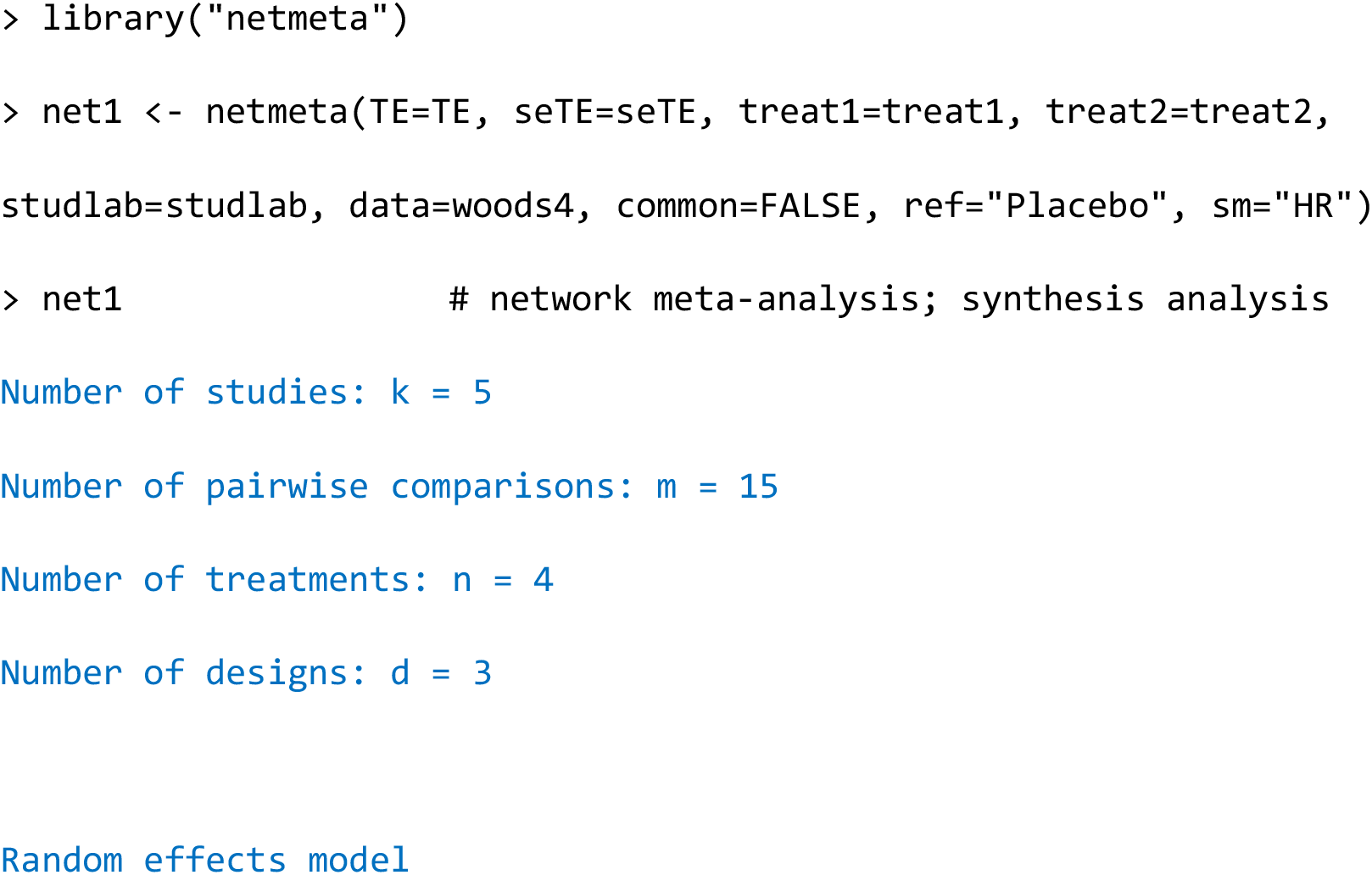
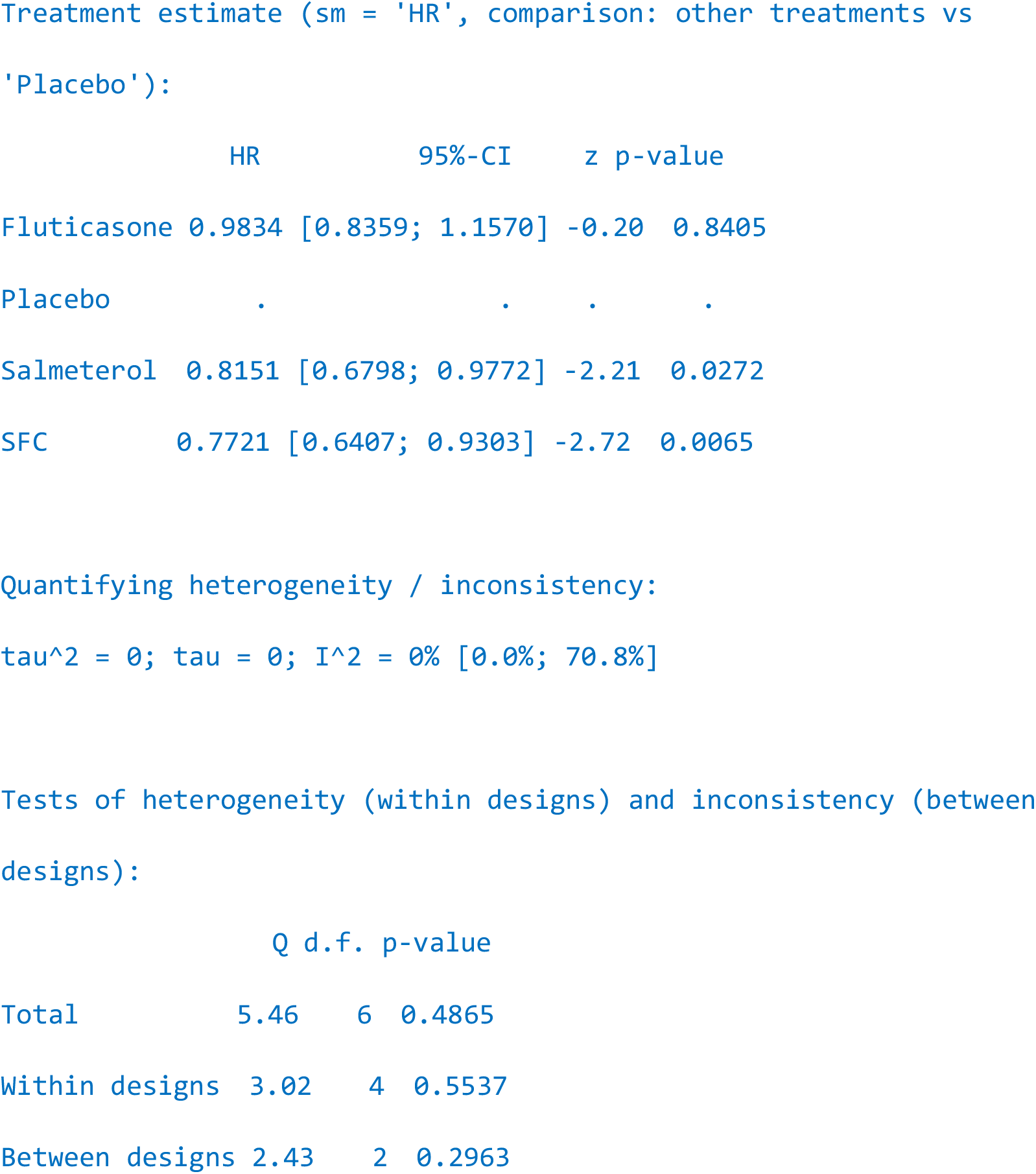

The pooling results are compared with those by the Bayesian approach of Woods et al. ^1^ in Table 3. Since the *τ* estimate of the REML method was 0, only the pooling results of the fixed-effect models are presented. We can confirm that the results are very similar between the two fixed-effect models; although the random-effects models often provide quite different results because of the differences of the principles of statistical inferences. Actually, the *τ* estimate reported by the Bayesian approach of Woods et al. ^1^ was 0.36; however, these differences are simply explained by the principles of the two different methodologies ^18,19^. The frequentist validities of the REML-based inference methods are shown in many previous reports ^12,20^.

**Table 3.**
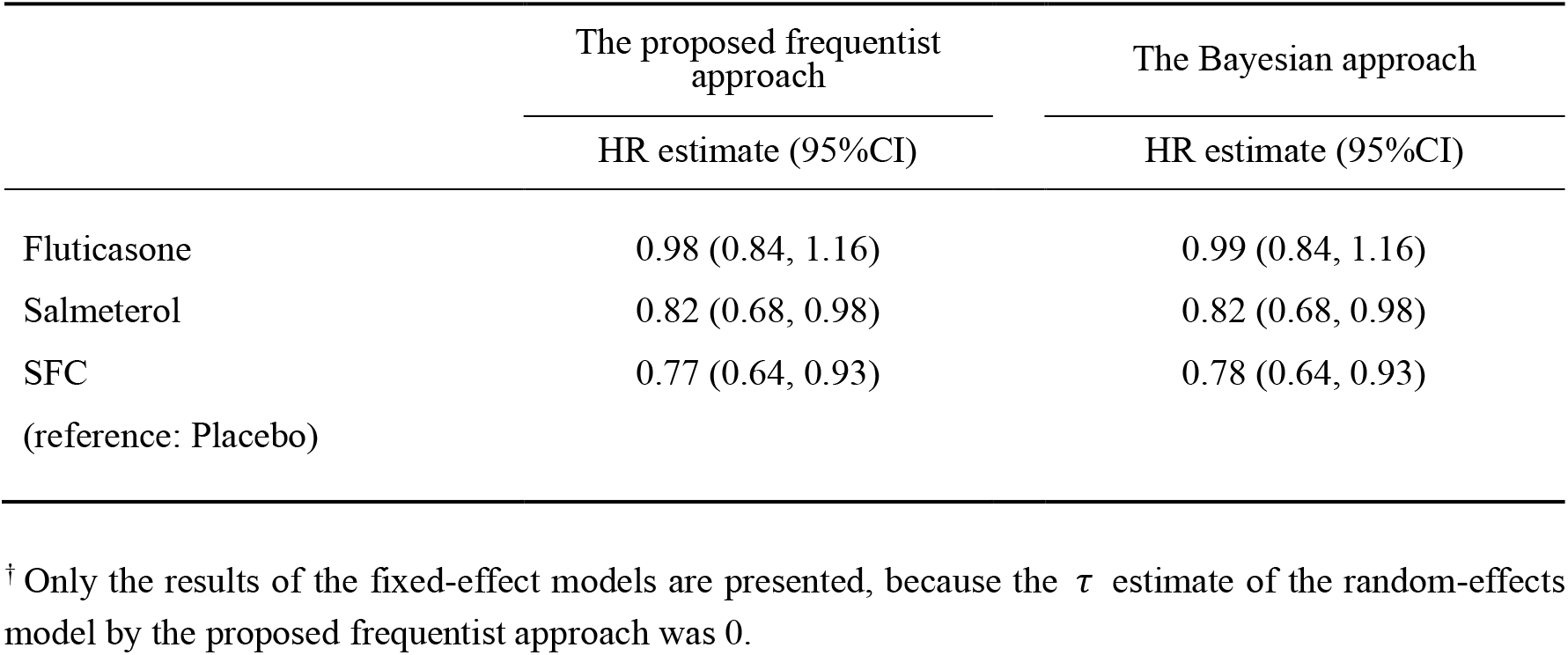
Results of the network meta-analyses of COPD by the proposed frequentist approach and the Bayesian approach (the latter data was cited from Woods et al. ^1^)^†^.

## Data Availability

The network meta-analysis data are available in Woods et al. (2010; BMC Med Res Methodol 10:54).

## Example R code

Example R code is available at https://www.ism.ac.jp/∼noma/example_survNMA.r

## Acknowledgements

This study was supported by Grants-in-Aid for Scientific Research from the Japan Society for the Promotion of Science (grant numbers: JP23K11931, JP22H03554, and JP24K21306).

